# Social and mental health risks faced by undocumented migrants during the COVID-19 pandemic: Evidence from three surveys in France

**DOI:** 10.1101/2021.10.14.21264999

**Authors:** Gosselin Anne, Longchamps Cécile, Oulahal Rachid, Derluyn Ilse, Ducarroz Simon, Skovdal Morten, Verelst An, Sturm Gesine, Desgrées du Loû Annabel, Melchior Maria

**Author notes:** **Corresponding author :** Anne Gosselin, Mortality, Health, Epidemiology Unit, French Institute for Demographic Studies (Ined) 9 cours des Humanités, 93322 Aubervilliers, France, |.

## Abstract

**Background:** The often-precarious life circumstances of undocumented migrants are likely to heighten the detrimental impact of the COVID-19 pandemic on their lives. Given the paucity of research exploring how undocumented migrants are affected by the COVID-19 pandemic, we set out to explore the association between being an undocumented migrant and a range of social and mental health measures.

**Methods:** Our study draws on three complementary surveys conducted among migrants in France between April 1^st^ and June 7^th^ 2020 (APART TOGETHER, MAKASI, ECHO; n = 716). We tested associations between eight outcome measures, covering health literacy, prevention behaviours, perceptions of government responses, livelihoods and mental health (PHQ-9 score), and the participants’ legal status as either undocumented or documented. We modelled the probability of food insecurity increase, job loss, depression, and responses to SARS-COV-2 symptoms with logistic regression models, adjusted for age, gender and legal status.

**Results:** Undocumented migrants had a higher probability of experiencing food insecurity increase (aORs=10.40 [3.59, 30.16], and 2.19 [1.39, 3.50] in APART TOGETHER and ECHO), a higher probability of depression (aOR=2.65 [1.01, 6.97] in MAKASI). In all three surveys, undocumented migrants were more likely to lose their job (aORs=6.51 [1.18, 36.00], 8.36 [1.08, 64.70] and 3.96 [1.79, 9.16] in APART TOGETHER, MAKASI and ECHO respectively).

**Conclusion:** Our results suggest that the lives of undocumented migrants have been dramatically worsened by the COVID-19 pandemic, exposing and amplifying the inequalities facing this group. There is an urgent need for action to address these inequalities.

**Highlights**

- Undocumented migrants in Europe are often living in difficult socioeconomic circumstances and experience important barriers in access to healthcare
- Undocumented migrants are a hard-to-reach population and may be not included in surveys that explore the social and sanitary consequences of the COVID-19 pandemic
- To date there is scarce quantitative evidence on the social and mental health risk among undocumented migrants during the COVID-19 pandemic
- Our study shows that in France, undocumented migrants had a higher probability of losing their job, experiencing food insecurity increase and depression during the first wave of the Covid-19 pandemic (March-May 2020) than migrants with legal resident permit.
- Results on COVID-19-prevention knowledge are ambivalent and show area for improvement

## Introduction

Shortly after the upsurge of the COVID-19 pandemic, public health experts alerted the scientific community about the potential consequences of the pandemic on undocumented migrants. Undocumented migrants are here defined as persons who have no legal residence permit in France, and no current official claim for asylum or another legal status. The absence of a solid bond of trust with the statutory authorities, the lack of established means for the communication of health information, their frequently overcrowded living conditions and limited rights to health care have been identified as possible mechanisms explaining a higher burden of the COVID-19 pandemic on this group (1,2). However, quantitative evidence on this topic remains scarce and our study will be the first to empirically explore the associations between legal status and social and health outcomes during the pandemic.

In France, the estimated number of undocumented migrants is approximately 300,000 (3). Recent police evictions of camps in the Greater Paris area have overtly revealed the extreme vulnerability of undocumented migrants. However, not all undocumented migrants live in camps, temporary shelters or are homeless: the experience of being undocumented also affects a larger population of migrants living in variable situations with regards to housing and work. The median time it takes migrants in France to obtain a first residence permit of at least one year has estimated to be 4 years for men and 3 years for women among a random sample of persons from sub-Saharan Africa (4). During the period of absence of a resident permit, it is difficult for individuals to establish stable and health-promoting living conditions, obtain stable work, and access healthcare services (5–8), despite undocumented migrants’ entitlement to access free healthcare.

In order to control the COVID-19 pandemic, the French government ordered a lockdown between the 17^th^ of March and the 11^th^ of May 2020. Everyone was supposed to stay at home, except a limited selection of key workers. The population had to justify via a certificate (“attestation sur l’honneur”) each occasion on which they left their home, and only certain pre-specified reasons (e.g., food shopping, medical appointment) were allowed. Long-term shelters (i.e. hosting people for more than one night) were specifically settled during the lockdown for homeless persons. The data available in the general population suggest that the epidemic had a major impact on the economic activity and the population’s mental health (9), with important social inequalities (10). These negative consequences of the epidemic could have been particularly important for undocumented migrants, given their precarious living conditions, however to date there are few data on this topic.

Our study has therefore two objectives: first, to investigate the knowledge of COVID-19 prevention measures amongst undocumented migrants living in France, as well as their responses to COVID-19 symptoms; second, to examine whether being undocumented during the COVID-19 pandemic is associated with heightened food insecurity, job loss, and poor mental health.

## Methods

The study is based on three complementary survey conducted in France amongst migrants between April 1^st^ and June 7^th^ 2020, corresponding to the first wave of the COVID-19 epidemic and the associated lockdown.

APART TOGETHER is a cross-sectional study designed to explore the impact of the COVID-19 and social distancing measures on the psychological and social wellbeing of migrants and refugees, first in several European countries (Belgium, Denmark, France, Italy, Greece, Portugal, Spain, the UK, and Sweden) and then, in collaboration with WHO, upscaled worldwide. The study was conducted from 21st April to 30th October 2020 through an online survey available in 37 languages (French, English, Arabic, Turkish, Yoruba, Pashto …). 28,000 refugees and migrants living in 193 different countries and originating from 184 countries participated in the study (11). In the present study, we included participants who accessed the survey in France between 21 April and 7 June 2020, corresponding to 141 individuals (39.9% of persons who accessed the survey from France).

MAKASI is a community-based interventional study, which includes migrants from sub-Saharan Africa in the Greater Paris area experiencing social disadvantage, to assess the effectiveness of an empowerment intervention on risks in terms of sexual health. The research protocol includes a baseline questionnaire on participants’ socioeconomic circumstances and mental health, as well as follow-up questionnaires 3 and 6 months after intervention delivery. Questionnaires were completed in French or in English. The study was registered with Clinical Trials: NCT04468724 (12). At the time of the first lockdown and owing to the established protocol, we continued this follow-up by phone and added questions specific to the COVID-19 pandemic and lockdown situation. A total of 100 participants answered the study questionnaire and could be included in the statistical analyses for this study.

ECHO is a cross-sectional study conducted among residents of 18 short and long-term homeless shelters in the Paris (n=12), Lyon (n=5) and in Strasbourg (n=1) regions. The study interview, aiming to measure participants’ level of information regarding COVID-19, their knowledge, implementation and acceptability of various preventive measures, was conducted between May 2^nd^ and June 7, 2020, by trained interviewers, in person or by telephone. The questionnaire was administered in French, in English or in the participant’s language: 25% of questionnaires were completed with the assistance of a trained translator contacted by telephone (most frequently in Arabic, Pashto, Dari, Tigrinya, and Amharic). In total, 929 residents were invited to take part in the survey, 131 refused participation, 263 were unavailable, and 535 (57.6%) completed the study questionnaire (13).

## Ethical approvals

The APART TOGETHER study obtained the approval of the Ethics Committee of the Faculty of Psychology and Educational Sciences of Ghent University (2020-41). The survey protocol, questionnaire and informed consent form were also approved by the WHO Ethics Review Committee. Additionally, an approval from the Ethics Committee of the University of Toulouse was obtained for the French part of the study (CER-2020-271).

The MAKASI study was approved both by the Comité de Protection des Personnes Sud-Ouest et Outre-Mer (ID RCB 2018-A02129-46) and by the CNIL (Commission Nationale Informatique et Libertés), n°2215270 in France.

The ECHO study received approval of the Ethical Research Committee of the University of Paris (CER-2020-41).

## Measures

### Level of knowledge of COVID-19 and preventive measures

This dimension was ascertained by 5 items: knowledge about the asymptomatic transmission of the SARS-CoV-2 virus; sources of information on COVID-19; practice of preventive gestures; reaction towards symptoms; level of satisfaction with the information provided by the government.

### Living conditions and health during the crisis

This dimension was ascertained by three items: food insecurity increase, job loss since the lockdown, depression (PHQ-9 score).

## Statistical analyses

In the three studies, we used data collected between the 1st of April and 7th of June 2020. To test the association between participants’ legal status and their living circumstances as well as level of knowledge of COVID-19 and practice of preventive measures, we first described the sociodemographic characteristics of the three study samples, and we then tested associations between the eight outcomes and participants’ legal status (undocumented or not) using chi-square tests. Thirdly, we modelled the probability of food insecurity increase, job loss, depression, and responses to SARS-COV-2 symptoms with logistic regression models, adjusting for age, gender and legal status. All analyses were stratified by survey and conducted using SPSS (APART TOGETHER), R (ECHO) and STATA (MAKASI).

## Results

Research participants were mostly male, with a median age just above thirty years, and had a median duration of stay in France of about two years. Despite these similarities, the description of the population shows different profiles across the three surveys, with a gradient of disadvantage: the participants of APART TOGETHER mostly had a high level of education (55%); 27% were undocumented and a majority lived in their own home or were hosted by family or friends (72%); 24% had a job before the lockdown. The population of MAKASI was in an intermediate situation: 68% were undocumented but a majority (82%) lived in their own home/were hosted; 38% had a job before the lockdown. All of the ECHO study participants lived in long-term homeless shelters and 74% of them are undocumented; 26% had a job before the lockdown.

Comparing participants with a legal status to those who were undocumented showed that legal status was not associated with certain outcomes: the level of knowledge of asymptomatic transmission of the SARS-COV-2 virus which was generally low (between 1/3 and 1/5) and the satisfaction regarding official information which was relatively high, above 60% in all the subgroups (Table 2).

**Table 1.**
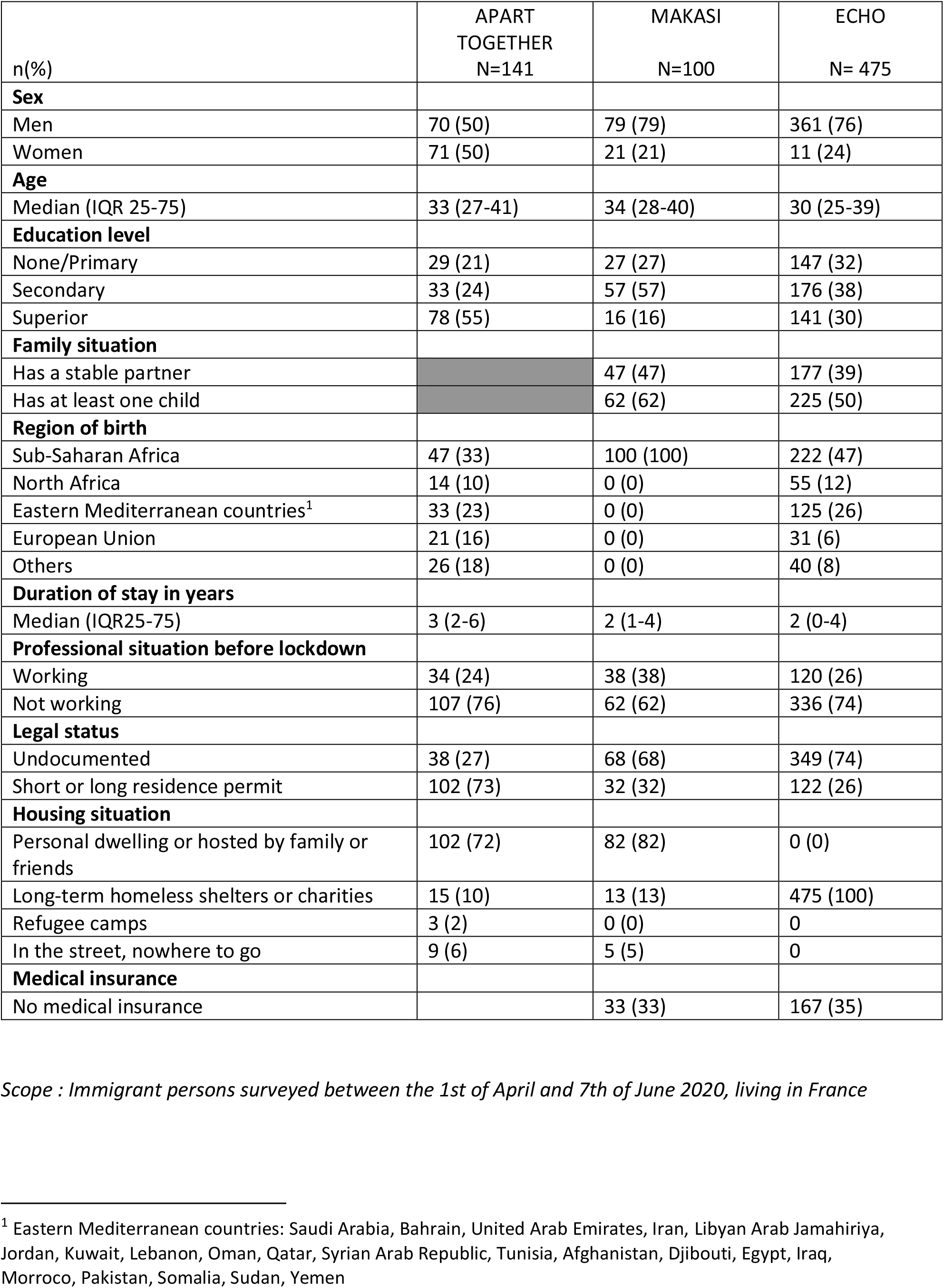
Sociodemographic characteristics of participants in APART TOGETHER, ECHO and MAKASI, immigrants in France April-June 2020.

| n(%) | APART<br>TOGETHER<br>N=141 | MAKASI<br>N=100 | ECHO<br>N= 475 |
| --- | --- | --- | --- |
| **Sex** |  |  |  |
| Men | 70 (50) | 79 (79) | 361 (76) |
| Women | 71 (50) | 21 (21) | 11 (24) |
| **Age** |  |  |  |
| Median (IQR 25-75) | 33 (27-41) | 34 (28-40) | 30 (25-39) |
| **Education level** |  |  |  |
| None/Primary | 29 (21) | 27 (27) | 147 (32) |
| Secondary | 33 (24) | 57 (57) | 176 (38) |
| Superior | 78 (55) | 16 (16) | 141 (30) |
| **Family situation** |  |  |  |
| Has a stable partner |  | 47 (47) | 177 (39) |
| Has at least one child |  | 62 (62) | 225 (50) |
| **Region of birth** |  |  |  |
| Sub-Saharan Africa | 47 (33) | 100 (100) | 222 (47) |
| North Africa | 14 (10) | 0 (0) | 55 (12) |
| Eastern Mediterranean countries<sup>1</sup> | 33 (23) | 0 (0) | 125 (26) |
| European Union | 21 (16) | 0 (0) | 31 (6) |
| Others | 26 (18) | 0 (0) | 40 (8) |
| **Duration of stay in years** |  |  |  |
| Median (IQR25-75) | 3 (2-6) | 2 (1-4) | 2 (0-4) |
| **Professional situation before lockdown** |  |  |  |
| Working | 34 (24) | 38 (38) | 120 (26) |
| Not working | 107 (76) | 62 (62) | 336 (74) |
| **Legal status** |  |  |  |
| Undocumented | 38 (27) | 68 (68) | 349 (74) |
| Short or long residence permit | 102 (73) | 32 (32) | 122 (26) |
| **Housing situation** |  |  |  |
| Personal dwelling or hosted by family or friends | 102 (72) | 82 (82) | 0 (0) |
| Long-term homeless shelters or charities | 15 (10) | 13 (13) | 475 (100) |
| Refugee camps | 3 (2) | 0 (0) | 0 |
| In the street, nowhere to go | 9 (6) | 5 (5) | 0 |
| **Medical insurance** |  |  |  |
| No medical insurance |  | 33 (33) | 167 (35) |

*Scope : Immigrant persons surveyed between the 1st of April and 7th of June 2020, living in France*
<sup>1</sup> Eastern Mediterranean countries: Saudi Arabia, Bahrain, United Arab Emirates, Iran, Libyan Arab Jamahiriya, Jordan, Kuwait, Lebanon, Oman, Qatar, Syrian Arab Republic, Tunisia, Afghanistan, Djibouti, Egypt, Iraq, Morocco, Pakistan, Somalia, Sudan, Yemen

**Table 2.** Level of knowledge about Covid-19, living conditions and mental health according to legal status, participants in APART TOGETHER, ECHO and MAKASI, immigrants in France April-June 2020.

|  | APART TOGETHER |  |  | MAKASI |  |  | ECHO |  |  |
| --- | --- | --- | --- | --- | --- | --- | --- | --- | --- |
|  | Migrants with resident permit | Undocumented migrants | p | Migrants with resident permit | Undocumented migrants | p | Migrants with resident permit | Undocumented migrants | p |
| **To know about the asymptomatic transmission** |  |  |  |  |  |  |  |  |  |
| Yes | - | - |  | 13 (42) | 38 (56) | ns | 85 (70) | 219 (64) | ns |
| No | - | - |  | 12 (39) | 17 (25) |  | 24 (20) | 75 (22) |  |
| Does not know | - | - |  | 6 (19) | 13 (19) |  | 13 (11) | 50 (15) |  |
| **Sources of information on Covid 19** |  |  |  |  |  |  |  |  |  |
| Posters in the streets |  |  |  | 3 (10) | 8 (12) | ns | 71 (62) | 201 (61) | ns |
| Television |  |  |  | 24 (77) | 47 (70) | ns | 88 (73) | 235 (69) | ns |
| Radio |  |  |  | 1 (3) | 6 (9) | ns | 44 (38) | 73 (22) | *** |
| Social networks : facebook, whatsapp | 49 (60) | 11 (30) | *** | 18 (58) | 39 (58) | ns | 100 (82) | 276 (81) | ns |
| Websites |  |  |  | 6 (19) | 20 (30) | ns | 89 (76) | 221 (67) | * |
| Word of mouth | 34 (41) | 7 (19) | ** | 5 (16) | 13 (19) | ns | 96 (83) | 247 (74) | * |
| Other |  |  |  | 2 (6) | 5 (7) | ns | 104 (88) | 282 (84) | ns |
| **Practiced prevention gestures** |  |  |  |  |  |  |  |  |  |
| Washing hands/hydroalcoholic gel | 74 (95) | 30 (88) | ns | - | - |  | 119 (98) | 329 (96) | ns |
| More physical distancing | 74 (95) | 29 (85) | * | - | - |  | 114 (93) | 322 (94) | ns |
| Cover nose and mouth in public | 65 (84) | 28 (82) | ns | - | - |  | 118 (98) | 327 (96) | ns |
| Avoid public transportation | 68 (89) | 25 (73) | ** | - | - |  | 98 (80) | 263 (78) | ns |
| Avoid going out | 72 (93) | 26 (76) | *** | - | - |  |  |  |  |
| **Response to SARS-COV-2 symptoms<sup>1</sup>** |  |  |  |  |  |  |  |  |  |
| Would go to a doctor or social worker in case of symptoms | 72 (92) | 27 (77) | ** | - | - |  | 99 (81) | 296 (86) | ns |
| Satisfaction with the diffusion of information by the government |  |  |  |  |  |  |  |  |  |
| Yes | - | - |  | 8 (26) | 18 (26) | ns | 73 (60) | 185 (54) | ns |
| Yes more or less | - | - |  | 11 (35) | 29 (43) |  | 19 (16) | 20 (20) |  |
| Not really | - | - |  | 5 (16) | 12 (18) |  | 7 (6) | 34 (10) |  |
| Not at all | - | - |  | 5 (16) | 8 (12) |  | 13 (11) | 32 (9) |  |
| Does not know |  |  |  | 2(6) | 1 (1) |  |  |  |  |
| Food insecurity increase | 11 (17) | 21 (63) | *** | 15 (48) | 45 (66) | * | 32 (27) | 150 (44) | *** |
| Job loss since the lockdown | 2 (2) | 5 (13) | *** | 4 (50) | 22 (73) | ns | 18 (38) | 49 (68) | *** |
| Depression(PHQ9, significant symptoms, score >10) | - | - |  | 18 (58) | 79 (79) | ** | 24 (22) | 97 (31) | * |

*p* : *p*-value chi2 test comparing persons with legal status and undocumented; \**p*<0.10; \*\**p*<0.05 \*\*\**p*<0.01; ns
1. In *Apart Together*, the question asked was as follows: In case I or one of my family members develops symptoms, I would contact a doctor or health care provider? Yes/No. In *ECHO*, the question was asked as follows: If you had symptoms of coronavirus, what would you do first: try and heal myself alone with herbal tea/plants, try to heal myself with medications, talk to social worker in a centre, call the 15, go to hospital, go to a doctor, other.

**Tableau 3.** Determinants of increased food insecurity, moderate to severe depression, job loss and attitude in case of symptoms, (logistic regressions adjusted on sex, age, and legal status).

|  | **APART TOGETHER**<br>**N=141** | **MAKASI**<br>**N=100** | **ECHO**<br>**N=475** |
| --- | --- | --- | --- |
|  | aOR [IC95%] | aOR [IC95%] | aOR [IC95%] |
| **Increased food insecurity** |  |  |  |
| Women (ref men) | 0,74 [0.27, 2.02] | 1.01 [0.36, 2.85] | 0.80 [0.50, 1.25] |
| 18-29 years (ref 30-39) | 0,35 [0.11, 1.15] | 1.05 [0.38, 2.88] | 1.13 [0.72, 1.77] |
| 40+ years (ref 30-39) | 0,39 [0.10, 1.47] | 0.54 [0.19, 1.53] | 0.87 [0.51, 1.49] |
| Undocumented migrant (ref migrants with legal status or nationality) | **10,40 [3.59, 30.16]** | 1.97 [0.81, 4.81] | **2.19 [1.39, 3.50]** |
| **Moderate to severe depression (PHQ9>=10)** |  |  |  |
| Women (ref men) |  | 4.29 [0.90, 20.43] | 1.54 [0.93, 2.54] |
| 18-29 years (ref 30-39) |  | 1.00 [0.33, 3.10] | 1.67 [1.01, 2.80] |
| 40+ years (ref 30-39) |  | 0.92 [0.29, 2.93] | 0.78 [0.41, 1.47] |
| Undocumented migrant (ref migrants with legal status or nationality) |  | **2.65 [1.01, 6.97]** | 1.60 [0.97, 2.73] |
| **Job loss among persons employed before lockdown** |  |  |  |
| Women (ref men) | 1,25 [0.25, 6.09] | **0.08 [0.00, 0.75]** | 0.34 [0.12, 0.96] |
| 18-29 years (ref 30-39) | 1,25 [0.21, 7.63] | 3.10 [0.32, 29.54] | 0.68 [0.26, 1.70] |
| 40+ years (ref 30-39) | 0,57 [0.05, 6.91] | 0.79 [0.09, 6.78] | 0.64 [0.22, 1.80] |
| Undocumented migrant (ref migrants with legal status or nationality) | **6,51 [1.18, 36.00]** | **8.36 [1.08, 64.70]** | **3.96 [1.79, 9.16]** |
| **To go and see a doctor/a social worker in case of symptoms** |  |  |  |
| Women (ref men) | 0,53 [0.16, 1.76] |  | 0.78 [0.44, 1.39] |
| 18-29 years (ref 30-39) | 0,74 [0.16, 3.34] |  | 1,51 [0.84, 2.69] |
| 40+ years (ref 30-39) | 0,53 [0.11, 2.52] |  | 1,41 [0.72, 2.83] |
| Undocumented migrant (ref migrants with legal status or nationality) | **0,28 [0.09, 0.92]** |  | 1,40 [0.80, 2.40] |

Yet, our results showed that migrants with a legal status had more information about the COVID-19 infection through various sources: websites, radio, social networks, as opposed to mainly television and street posters which were used to the same degree in both groups. The adherence to preventive measures was high, in general above 80% for all recommended measures. They were no differences between the two groups regarding washing hands, physical distancing and cover nose and mouth in public. The only difference we found between undocumented migrants and the others had to do with preventive measures that participants could not necessarily control if they kept on working (e.g. in APART TOGETHER: avoidance of public transportation: 73% in undocumented migrants vs. 89% among migrants with legal status, p<0.05). Undocumented migrants reported less frequently that they would see a doctor or a social worker in case of SARS-COV-2 symptoms in APART TOGETHER (77%). They had higher levels of food insecurity increase in all three surveys (between 44% and 66%), and experienced more job loss and a higher proportion of significant symptoms of depression than migrants who were documented, although for this last outcome, the observed prevalence rates differed greatly across populations: 79% in MAKASI and 31% in ECHO.

When adjusting for age and gender, certain differences between migrants with legal residence and undocumented migrants remained: the probability of experiencing food insecurity increase was higher among the undocumented (aORs=10.40 [3.59, 30.16], and 2.19 [1.39, 3.50] in APART TOGETHER and ECHO), as was the probability of depression in MAKASI (aOR=2.65 [1.01, 6.97]). In contrast, undocumented migrants had a decreased probability of seeing a doctor in case of symptoms in APART TOGETHER (aOR=0.28 [0.09, 0.92]). In all three surveys, amongst persons who had a job before the lockdown, undocumented migrants were more likely to lose their job (aORs=6.51 [1.18, 36.00], 8.36 [1.08, 64.70] and 3.96 [1.79, 9.16] in APART TOGETHER, MAKASI and ECHO respectively).

## Discussion

This study brings together data from three surveys directed at migrants during the first COVID-19 lockdown in France.

Overall, the reported levels of social disadvantage are very high: food insecurity, job loss and feelings of depression were experienced by a large share of the undocumented migrant population compared to the general population (14). The situation regarding knowledge on prevention is ambivalent: on the one hand, we found high levels of adherence to preventive gestures, which were also reported in qualitative research (15,16). On the other hand, the knowledge of asymptomatic transmission of the SARS-COV-2 virus is rather low, which could mean the public communication was focused only on certain preventive gestures (physical distancing, washing hands) and not on delivering a full information about how the disease spreads.

This paper also shows that irrespective of the different surveys, migrants’ legal status appears to shape their experience of the pandemic and related lockdown. Undocumented migrants were consistently disadvantaged on all the outcomes we examined: not only did they appear to be more hit by the lockdown in terms of living circumstances and mental health, but it also seems that they were less inclined to see a doctor in case of symptoms. These social inequalities could contribute to undocumented migrants’ high levels of exposure to the virus and high morbidity and mortality once infected (17).

Finally, our study also shows that among undocumented migrants, all situations are not equivalent. Undocumented long-term homeless migrants (ECHO study) had more favourable indicators on several dimensions: they were more often aware of asymptomatic transmission than more settled migrants (MAKASI) (64% vs 56%), and their practice of prevention gestures was higher. Their level of depression was high, but lower than in MAKASI, corresponding to migrants in a more intermediate situation. Finally, the fact that they were undocumented did not seem to impact on their attitude in case of symptoms, suggesting that they would trust the system enough to go to the doctor if needed, as opposed to what is observed among the participants of APART TOGETHER. The fact that long-term homeless migrants were provided regular information by the social workers of the structure and that they could benefit from social and sanitary support during this period could explain these differences.

Our study has several limitations. First, the very attempt to compare different surveys is challenging, as all the questions were not necessarily asked in the same way and with the same answer modalities, which can lead to information bias. As an example, legal residence status was not asked the same way in all three surveys, and although all of them included the “undocumented” modality, all three did not collect information on participants’ resident permit in the same way, which prevented us from more precise analyses on the different administrative situations The MAKASI survey collected the information on the length of the legal permit (less than one year, 1y and more, 10years or more, Citizenship) whereas in ECHO, participants were asked whether they had a legal resident permit/an asylum claim. Second, although the total number of participants is 716, in each survey the number of individuals is relatively limited, precluding more in-depth analyses that would take more cofounders into account in multivariate analyses.

However, our study provides empirical data on a hard-to-reach population, namely undocumented migrants in France, and reports on their experiences of the first French lockdown and their knowledge of COVID-19. These results lead to several considerations in terms of public health campaigns and the fight against COVID-19. First, preventive messages should include not only specific instructions for prevention, but also key messages about the COVID-19 illness. Previous research based on ECHO data showed that according to the region of origin, the level of knowledge and practice of prevention gestures was different and this should probably be taken into account for designing tailored public health messages (13). Possibly, regarding vaccine campaigns, the sole instruction to get vaccinated is not sufficient and information on the ways in which the vaccine works and was tested could be key to ensure the success of future vaccination campaigns. Finally, migrants’ legal status is key with regard to their experience of the pandemic.

## Conclusion

Migrants are especially at risk in the COVID-19 pandemic, in relation with their living conditions and social situation (10,18). Our results suggest that the lack of legal residence permit can enhance many difficulties and heighten socioeconomic and mental health risks. From a research perspective, this means that more data needs to be collected with regards to country of birth, ethnicity and legal residence in the surveys investigating the socioeconomic and health impacts of the pandemic (19,20). Only with this data we will be able to understand the mechanisms that expose and amplify the risks experienced by undocumented migrants in France.

On the policy-level, our results suggest that specific interventions to improve the employment and housing situation of undocumented migrants are greatly needed, as well as programs that can enhance their access and trust in the healthcare system. In a broader perspective, many experts have advocated for guaranteeing access to a legal residence to undocumented migrants to limit the impact of the epidemic on their health, and our results definitely show how this recommendation is relevant in the current context.

## Data Availability

All data produced in the present work are contained in the manuscript

## Conflict of interest

None declared

## Funding information

The MAKASI study was supported by the French National Agency for research on AIDS and Viral hepatitis (ANRS) and Health regional agency (Agence régionale de santé Ile-de-France), as well as Université de Paris (ANR-18-IDEX-001).

The ECHO study was supported by l’Institut Convergences MIGRATIONS/CNRS (French Collaborative Institute on Migrations), (ANR-17-CONV-0001), by Santé Publique France and ANR Flash-Covid 19.

## Acknowledgements

**APART TOGETHER**. The Apart Together Sudy group in France included Gesine Sturm, Rachid Oulahal, Filipe Soto Galindo, Julia de Freitas Girardi and Yagmour Gökduman. The authors also thank the entire APART TOGETHER academic consortium, especially Ilse Derluyn, An Verelst, Eva Spiritus-Beerden and Morten Skovdal as well as the WHO for support and dissemination of the study. A special thanks to all participants as well as to our partners and professionals who helped to communicate the survey to participants of hard to reach groups.

**MAKASI**. The MAKASI Study Group included Annabel Desgrées du Loû, Nicolas Derche, Flore Gubert, Romain Mbiribindi, Maria Melchior (responsables scientifiques), Séverine Carillon, Virginie Comblon, Karna Coulibaly, Angèle Delbe, Jacques Ebongue, Ruth Foundje, Fabienne El Khoury, Charles Gaywahali, Anne Gosselin, Veroska Kohou, France Lert, Jean Lusilu-Voza, Belinda Lutonadio, Yves Nyemeck, Eve Plenel, Patricia Mbiribindi, Thierry Miatti, Jean-Paul Ngueya, Andrainolo Ravalihasy, Valéry Ridde, Jean-Noël Senne, Oumar Sissoko, Corinne Taéron, Faya Tess, Iris Zoumenou.

The authors would like to thank all the persons who participated in the Makasi study ; thanks to the peers committee for its support in preparing the questionnaire, AFRICASYS society for statistical support, Marie Soulié for communication tools, Emmanuel Lamotte for website. They also want to thank the members of the MAKASI Study Group:

We would also like to thank the NGO SOLTHIS who supported this research.

**ECHO**. The authors want to thank the ECHO scientific comittee includes Lisa Crouzet, Lionel Pourtau, Cécile Allaire, Anne-Claire Colleville and Tarik El Aarbaoui. The ECHO study group includes Nicolas Vignier, Pierre Anquetil, Léa Balage, Estelle Dussert, Betty Girard, Laure Luyinga Nzuzi, Hermine Metias, Nathalie Oprescu, Philippe Rebouffat-Roux (Habitat et Humanisme), Anahaid Armenian, Marianne Auffret, Perrine Leclerc (Association Aurore), Faouzi Bertrand (Groupe SOS), Mourad Bouderbal (Croix Rouge Française), Antoine Denis (SIAO 67), Christelle Witczak (Empreintes Sud 77), François Fortin (La Rose des Vents).

The authors would also like to thank all the persons who participated in the study.

## Notes

### Competing Interest Statement

The authors have declared no competing interest.

### Clinical Protocols

https://clinicaltrials.gov/ct2/show/NCT04468724

### Author Declarations

The APART TOGETHER study obtained the approval of the Ethics Committee of the Faculty of Psychology and Educational Sciences of Ghent University (2020-41). The survey protocol, questionnaire and informed consent form were also approved by the WHO Ethics Review Committee. Additionally, an approval from the Ethics Committee of the University of Toulouse was obtained for the French part of the study (CER-2020-271). The MAKASI study was approved both by the Comite de Protection des Personnes Sud-Ouest et Outre-Mer (ID RCB 2018-A02129-46) and by the CNIL (Commission Nationale Informatique et Libertes), nb 2215270 in France. The ECHO study received approval of the Ethical Research Committee of the University of Paris (CER-2020-41).

